# Beyond the Needle: Touch Activated Phlebotomy for Autism-Friendly Blood Sampling

**DOI:** 10.64898/2026.04.22.26351115

**Authors:** Alexander Cameron, Gabriella Rossetti, Teresa Tavassoli, David Field

## Abstract

**Purpose:** Blood draws have been associated with significant discomfort, especially for individuals with sensory hypersensitivity, as is common in autism. This results in avoidance of medical appointments and creates difficulties for scientific studies recruiting from this population. Touch Activated Phlebotomy (TAP) is a novel capillary blood collection technique that reduces the discomfort of blood draws, and here we aimed to assess its tolerability to autistic adults. Our secondary aim was to assess whether capillary and venous blood provide equivalent measurements of Vitamin B6 concentrations.

**Methods:** 23 participants (11 autistic: 12 non-autistic) were recruited, and two TAP devices were administered before providing pain ratings. Traditional venipuncture was also carried out in the non-autistic individuals, with the same pain measures reported. Enzyme Linked Immunosorbent Assays (ELISAs) were conducted to quantify concentrations of Vitamin B6.

**Results:** The TAP device caused significantly less pain than the traditional venipuncture procedure. Furthermore, TAP pain ratings in autistic individuals did not differ meaningfully from non-autistic individuals. Vitamin B6 concentrations showed minimal bias and good agreement between capillary and venous blood, and high repeatability between repeated capillary samples. No clear difference in Vitamin B6 concentrations was observed between autistic and non-autistic participants.

**Conclusion:** TAP is a well-tolerated method of obtaining capillary blood samples from autistic adults for medical and research purposes, and this has the potential to reduce avoidance of medical appointments in this population. Like most analytes tested to date, measurement of Vitamin B6 in capillary blood is a valid and reliable alternative to traditional venous samples.

## Introduction

Phlebotomy is one of the most common medical procedures in the United Kingdom, with approximately 100 million blood tests ordered by General Practitioners (GPs) each year (Watson et al., 2023). Highlighting the importance of blood draws for general health, Watson et al. (2023) reported that the most common reasons for blood test referral were symptom investigation (43.2%), monitoring of existing disease (30.1%), monitoring of an existing medication (10.1%), follow-up of previous anomaly (6.8%), and patient requested (1.5%). However, a highly prevalent challenge for blood drawing procedures is the associated fear. Approximately 20-30% of young adults have a fear of needles (McLenon & Rogers, 2019), with similar, if not higher, rates being observed in autistic individuals (Berner et al., 2024), leading to needle phobia being the most diagnosed specific phobia in autistic adults (Leyfer et al., 2006). This high rate of needle phobia may be explained by the high rates of sensory hypersensitivity in autistic individuals (Maclennan et al., 2022). The consequence of this fear is observable, with ∼52% of autistic individuals avoiding blood draws due to this fear and 95.5% reporting that pain is the most common reason for their fear of needles (Alsbrooks & Hoerauf, 2022). Furthermore, the high prevalence of fear and discomfort results in research studies avoiding blood draws in autistic individuals, leading to a significant lack of representation (Berner et al., 2024).

Beyond the challenges faced due to patient/participant fear, there are additional difficulties surrounding blood draws. One challenge, especially for research studies, is the extensive training required for phlebotomy, often requiring 40+ hours of classroom training along with 40+ hours of practical training, taking several months to complete (Warekois & Robinson, 2015).

Furthermore, risks to the patient/participant, including hematoma, thrombosis, infection, nerve damage, arterial puncture, and vasovagal reactions (Serra et al., 2018), make phlebotomy both a practical challenge for clinicians/researchers and a daunting experience for patients/participants.

Although alternative methods to phlebotomy exist, such as finger pricks, they often induce comparable discomfort and collect little blood, making sample analysis challenging (Serafin et al., 2020). In recent years, capillary blood collection devices have been developed with the aim of being less painful and self-administrable while still collecting an adequate amount of blood. The Touch Activated Phlebotomy (TAP) device has been developed by YourBioHealth (formerly Seventh Sense Biosystems) as one of these new techniques (Blicharz et al., 2018). The TAP device uses an array of microneedles, each smaller than an eyelash, which puncture the upper dermis for less than 250 milliseconds. Due to this fast and shallow penetration, the devices cause less pain than traditional venipuncture that requires much deeper penetration for longer durations.

YourBioHealth state that their TAP devices cause virtually no pain, with meta-analyses supporting this statement by showing that they cause significantly less pain than venipuncture procedures (Schröder et al., 2025). However, no studies have investigated whether this claim can be extended to autistic individuals. Furthermore, another remaining question is whether the blood collected from the capillaries is comparable to blood collected from venipuncture. Several studies have made progress in answering this question, as Catala et al. (2018) demonstrated that of 45 analytes measured, 39 showed a strong correlation between capillary and venous blood. Although this provides promising evidence for using capillary blood as an alternative measure for blood analyte concentrations, it is evidently analyte dependent. Therefore, a specific investigation into each analyte must be conducted before drawing direct comparisons between quantifications made from two types of blood. Here, for the first time, we will address this question for Vitamin B6.

The present study aims to answer three research questions. Firstly, how does the pain experienced during a phlebotomy blood draw compare with a TAP capillary blood draw? Secondly, is there a significant difference in the pain experienced during a TAP capillary blood draw between autistic and non-autistic individuals? Finally, can Vitamin B6 concentrations measured in capillary blood collected using a TAP device be used interchangeably with those measured in venous blood? Vitamin B6 has been chosen as the analyte of measurement for this study because, to the researcher’s best knowledge, no studies have compared the quantity of Vitamin B6 between capillary and venous blood. Analyte-by-analyte investigations are required when comparing analyte quantification performed using capillary and venous blood.

Furthermore, as this project aims to make blood drawing a more accessible method when conducting studies with autistic individuals, we have picked an analyte (Vitamin B6) in which the inactive forms have been identified to be significantly higher in autistic compared to non-autistic individuals (Zhang et al., 2024; Adams et al., 2006), whereas the biologically active form, pyridoxal-5-phosphate, has been reported to be deficient (Raiten et al., 1984). In the current study, the quantification technique used will measure the sum of all forms of Vitamin B6. It is expected that the autistic group will have a higher level of Vitamin B6. The second research aim of this project is to compare the tolerability of the TAP device blood draw between autistic and non-autistic individuals and assess the feasibility of recruiting autistic individuals to take part in studies that use the novel TAP blood collection technique.

Based on literature, we hypothesise that the TAP device will be a less painful technique than traditional venipuncture. Furthermore, we expect autistic individuals to find the TAP device more painful than non-autistic individuals, given their pre-existing discomfort with blood-drawing procedures. Moreover, based on the high proportion of analytes that show no significant difference in concentration between capillary and venous blood, we expect to see this same trend with Vitamin B6. Finally, based on existing literature, we expect total Vitamin B6 concentration to be higher in autistic than non-autistic participants.

## Methods

### Participants

A total of 23 participants were recruited from the local area surrounding the University of Reading, UK. The sample comprised 11 autistic participants (9 females, 2 males; mean age = 25.09 years, SD = 6.69) and 12 non□autistic participants (7 females, 5 males; mean age = 26.42 years, SD = 3.96). Autistic participants were all required to have had a formal diagnosis by a medical professional, while all non-autistic participants were screened for any diagnosed neurodevelopmental conditions. Any participants with known blood disorders or needle phobias were excluded. This study was reviewed and approved by the University of Reading Research Ethics Committee (UREC23_07). Participants provided informed consent at the start of the study.

### Design and Procedure

This study used a mixed design comparing two blood sampling methods: Touch Activated Phlebotomy Microselect model (TAP microselect) and phlebotomy. Three outcome measures were collected: Vitamin B6 concentration (ng/ml), expected pain of procedure (0-10 Likert scale, 0 = “No pain”, 10 = “Most intense pain imaginable”), actual pain of procedure (0-10 Likert scale, 0 = “No pain”, 10 = “Most intense pain imaginable”). The pain ratings were collected on paper with the researcher out of the room to avoid any influence this might have on responses.

For each of the TAP procedures, the site of blood extraction was below the shoulder on the outside of the arm. The site was cleansed using an alcohol swab before the device was administered. The TAP device was left on the individual’s arm for 2 minutes or until the collection tube was full (1000μL). This was repeated twice to obtain two TAP samples. The phlebotomy procedure was conducted by a trained phlebotomist using a 23-gauge butterfly needle, collecting 9ml of blood. For both procedures, blood was collected in a K2EDTA collection tube, then centrifuged at 1,000g for 15 minutes before the supernatant plasma was aliquoted and stored at –80°C with only one freeze-thaw cycle.

To assess participants pain ratings, they were first presented with information about the procedure (either TAP or phlebotomy) and then asked to rate how painful they expected the procedure to be. Following this, the procedure was conducted, and participants were asked to rate the actual pain of the procedure. This was repeated twice for TAP device administration. Only the non-autistic group experienced both phlebotomy and TAP administration due to feasibility concerns around the recruitment of autistic participants for phlebotomy. The order of TAP and phlebotomy was counterbalanced in the non-autistic group to balance order effects on pain ratings. Participants were reimbursed £10 for their time.

### Analysis

Total Vitamin B6 levels were measured as opposed to any of the individual B6 vitamers. Concentrations were measured using a commercially available Enzyme-Linked Immunosorbent Assay (ELISA) kit (Vitamin B6 ELISA Kit; Abbexa Ltd., Cambridge Science Park, Cambridge, UK; Cat. No. abx350884). All samples were run in duplicates and were analysed using ThermoFisher’s Multiskan SkyHigh Microplate Spectrometer (Thermo Scientific, Waltham, MA, USA). Absorbance was read at 450nm. Raw optical density (OD) values were first visually inspected for plate artefacts, outliers, and assay drift. The standard curve was generated using a 4-PL model, generated in R using the *drc* package. The parameters of the generated 4-PL model were used to interpolate raw sample OD values into concentrations based on the standards provided. No samples fell outside of the detectable range. Samples with intra□assay coefficients of variation (CVs) greater than 15% were excluded. No samples fell outside of the detectable range or exceeded a CV of 15%, thus none were excluded

Data was analysed and visualised using RStudio (RStudio version 2025.5.1.0, R version 4.2.2) using parametric and non-parametric statistical tests depending on the distribution of the data. Equal distribution was assessed using a Shapiro-Wilk test, and homogeneity of variance was then assessed using either a Levene’s test (if the data were normally distributed) or a Bartlett’s test (if the data were not normally distributed). Pain scores were compared between autistic and non-autistic participants using between-group analyses. Within the non-autistic group, paired analyses were conducted to compare pain experienced from the two blood collection techniques.

Agreement between venous blood collected by phlebotomy and capillary blood collected using the TAP device for Vitamin B6 measurement was assessed using Bland–Altman analysis (Bland & Altman, 1986; Bland & Altman, 1999), including regression of the differences on the mean to assess proportional bias. Reliability (repeatability) of Vitamin B6 measurements obtained from repeated TAP capillary sampling was assessed using Bland–Altman analysis, the Intraclass Correlation Coefficient (two-way mixed effects, absolute agreement; ICC(A,1)), and measures of precision including the Standard Error of Measurement (SEM) and Coefficient of Variation (CV%). Measures of agreement and reliability were interpreted in reference to previously published intra-individual biological variation for Vitamin B6 in healthy adults reported by European Federation of Clinical Chemistry and Laboratory Medicine biological variation database (Talwar et al., 2005, Aarsand et al., 2026). Finally, a between-group analysis was used to compare capillary Vitamin B6 concentrations between autistic and non-autistic participants.

## Results

### Descriptive statistics

Descriptive statistics for participants’ age, plasma Vitamin B6 concentration, and pain scores are reported in Table 1 for the autistic and non-autistic groups.

**Table 1:**
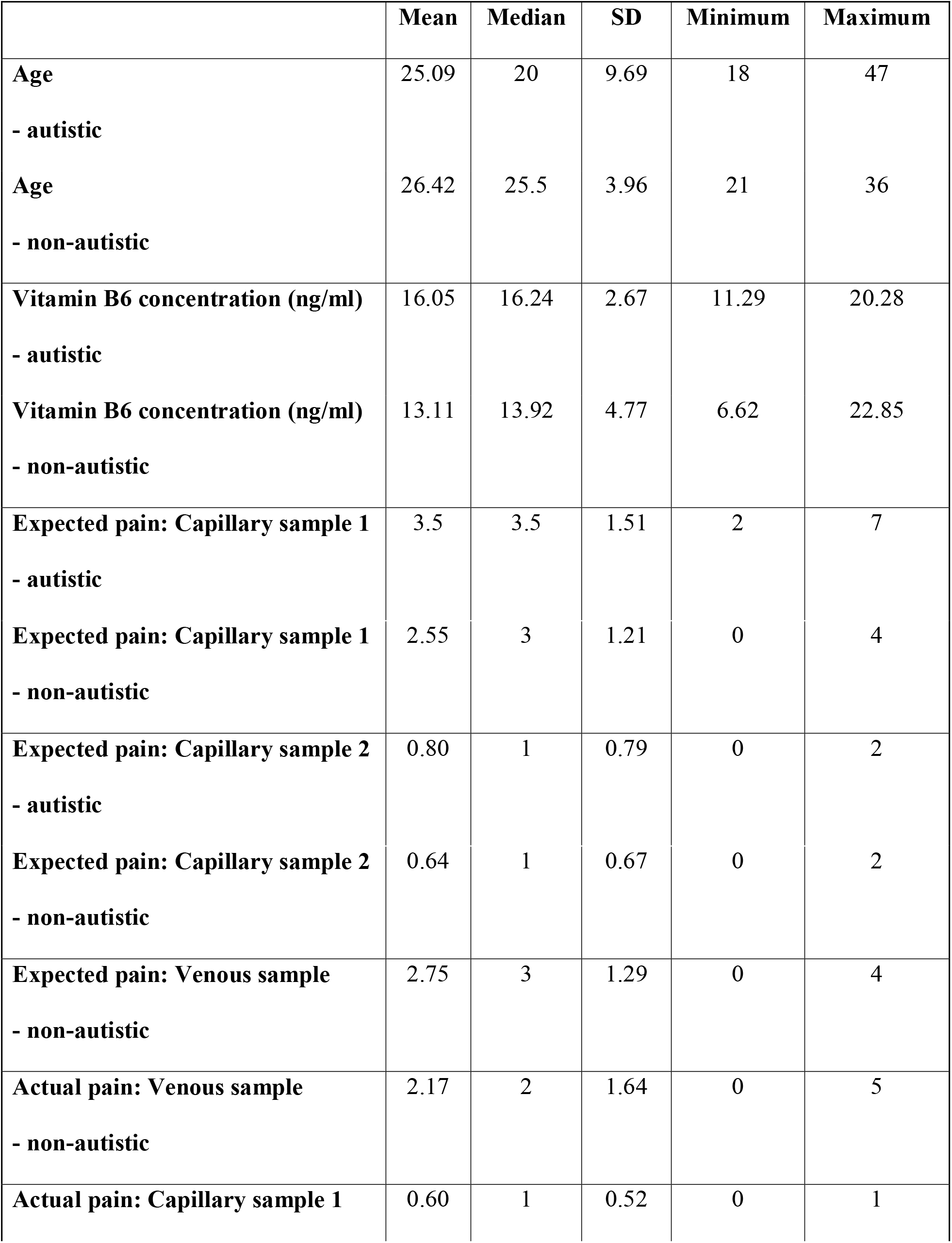
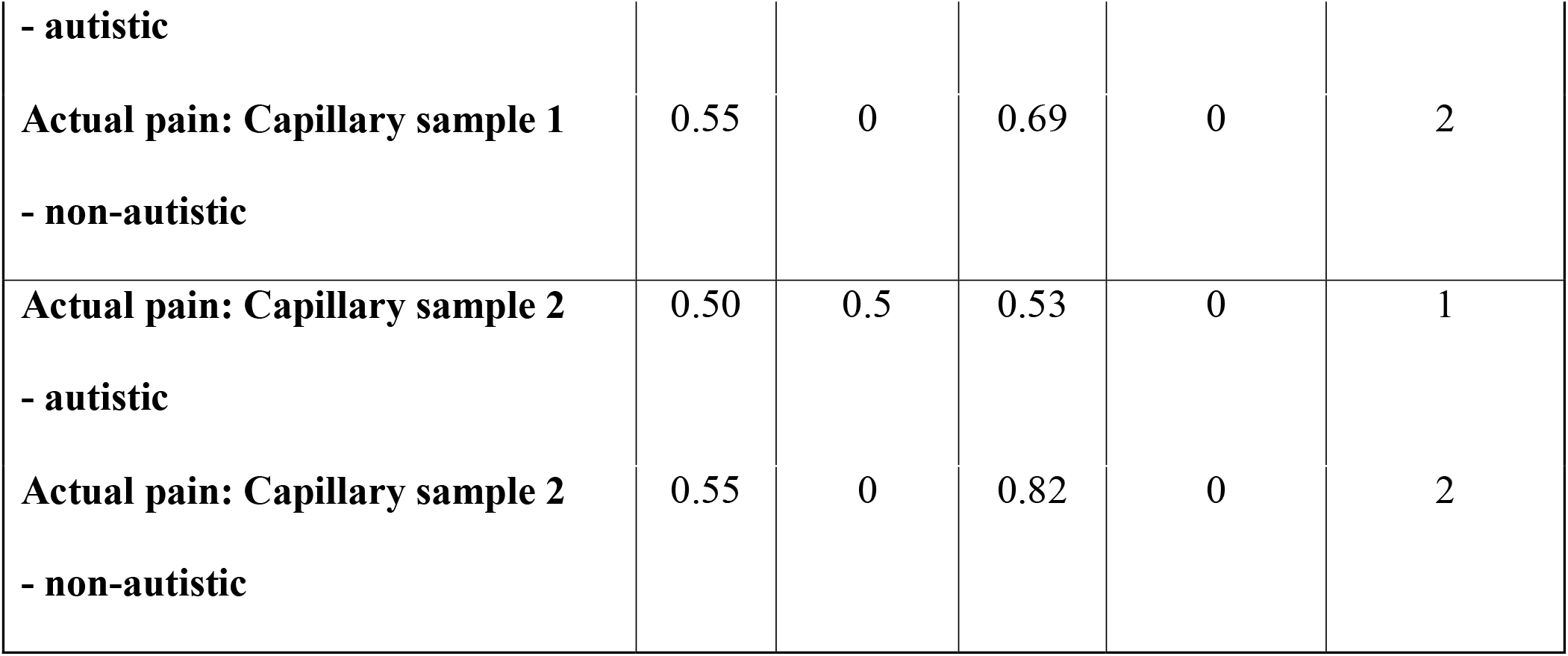

(Table 1)

### Comparing pain experienced during phlebotomy blood draws with TAP capillary blood draws

The assumption of homogeneity of variance was assessed using a Bartlett’s test, and the data were found to be heterogeneous in variance. Therefore, a paired Wilcoxon signed-rank test was conducted to compare the pain experienced from the phlebotomy procedure with the TAP procedure in the non-autistic group (Figure 1). The test showed that the TAP procedure caused significantly less pain than the phlebotomy procedure (V = 55, r = .59, p = .006).

**Figure 1.**
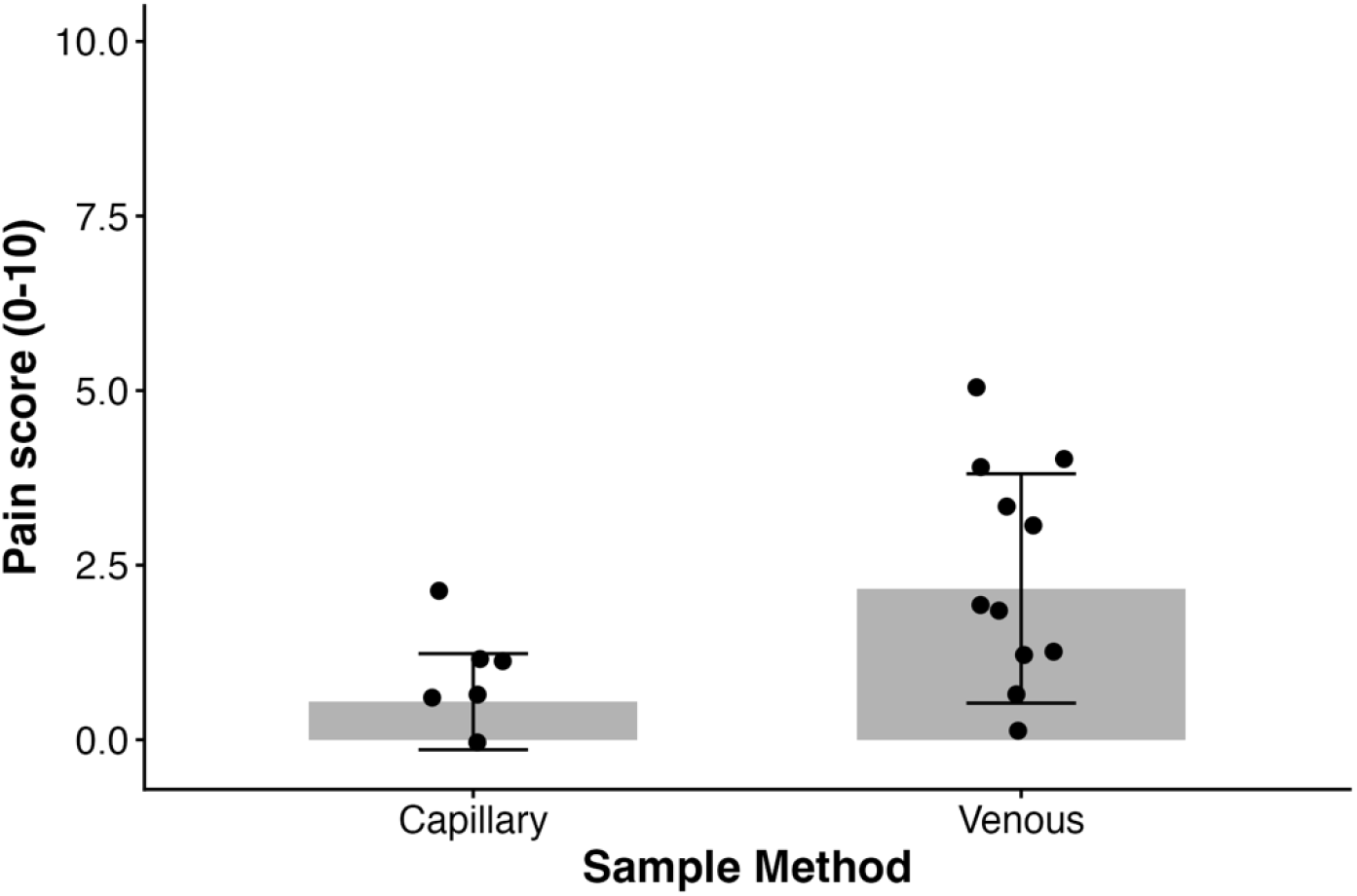
Comparison of pain between capillary and venous sampling. Comparing the pain experienced by non-autistic individuals when undergoing a TAP capillary blood sample compared to a venous phlebotomy sample. Error bars reflect standard deviation.

### Comparing expected and experienced pain scores between the autistic and non-autistic participants

Due to violated assumptions of normal distribution, a Mann-Whitney U (Wilcoxon rank sum) test was conducted to compare the expected pain experienced during the TAP procedure between the autistic and non-autistic participants (Figure 2, left panel). The same test was also conducted comparing actual pain from the TAP procedure between the autistic and non-autistic participants (Figure 2, right panel). This showed that there was no significant difference in mean expected pain experienced due to the TAP procedure between the autistic and non-autistic participants (W = 73, rank-biserial r = -.64, r = .29, p = .200). Furthermore, there was no statistically significant difference in mean experienced pain between the autistic and non-autistic groups (W = 59, rank-biserial r = -.93, r = .06, p = .795).

**Figure 2.**
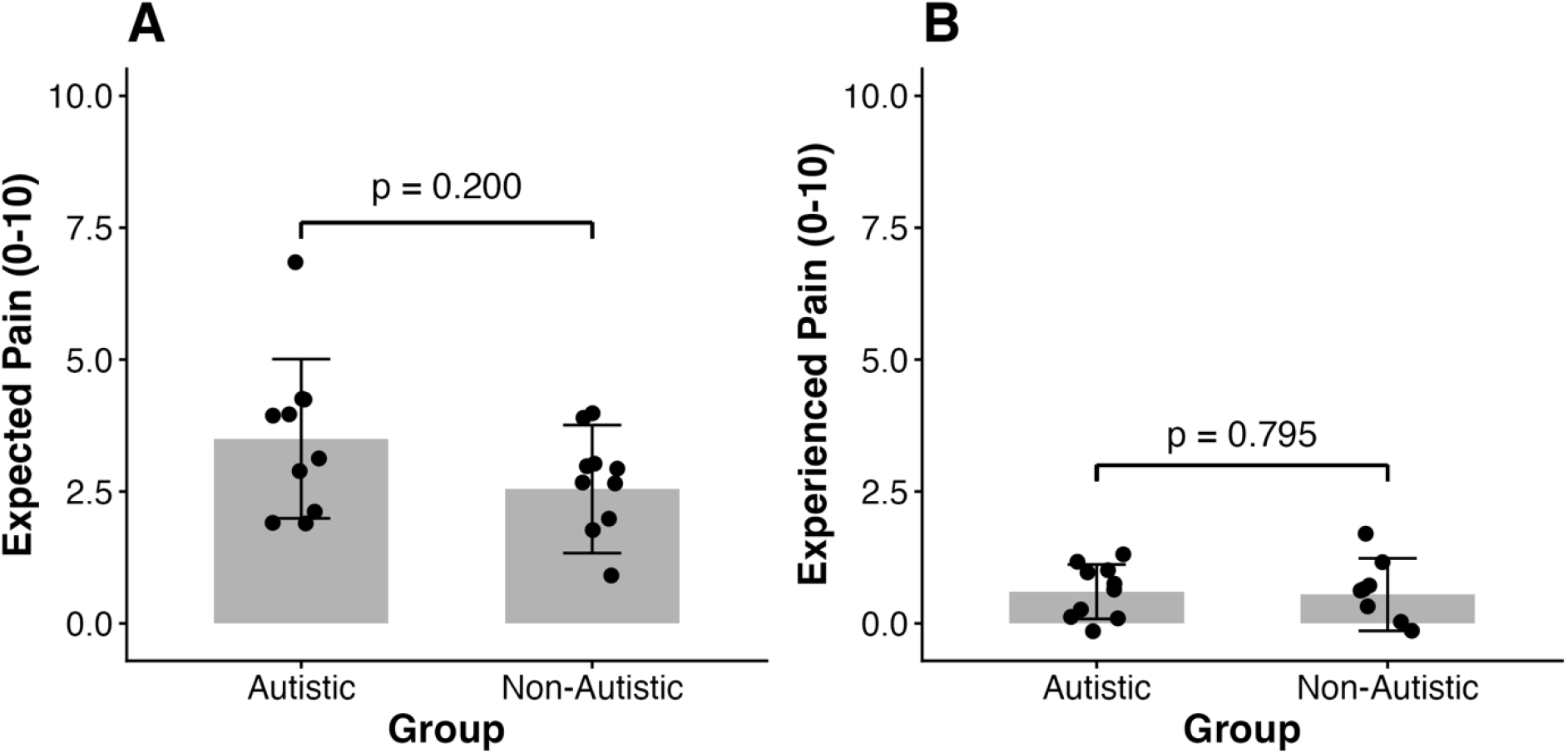
Comparison of pain between Autistic and Non-Autistic individuals (capillary samples only). (A) Pain expected and (B) Pain experienced by autistic and non-autistic individuals when undergoing a TAP capillary blood sample. Error bars reflect standard deviation.

**Figure 3.**
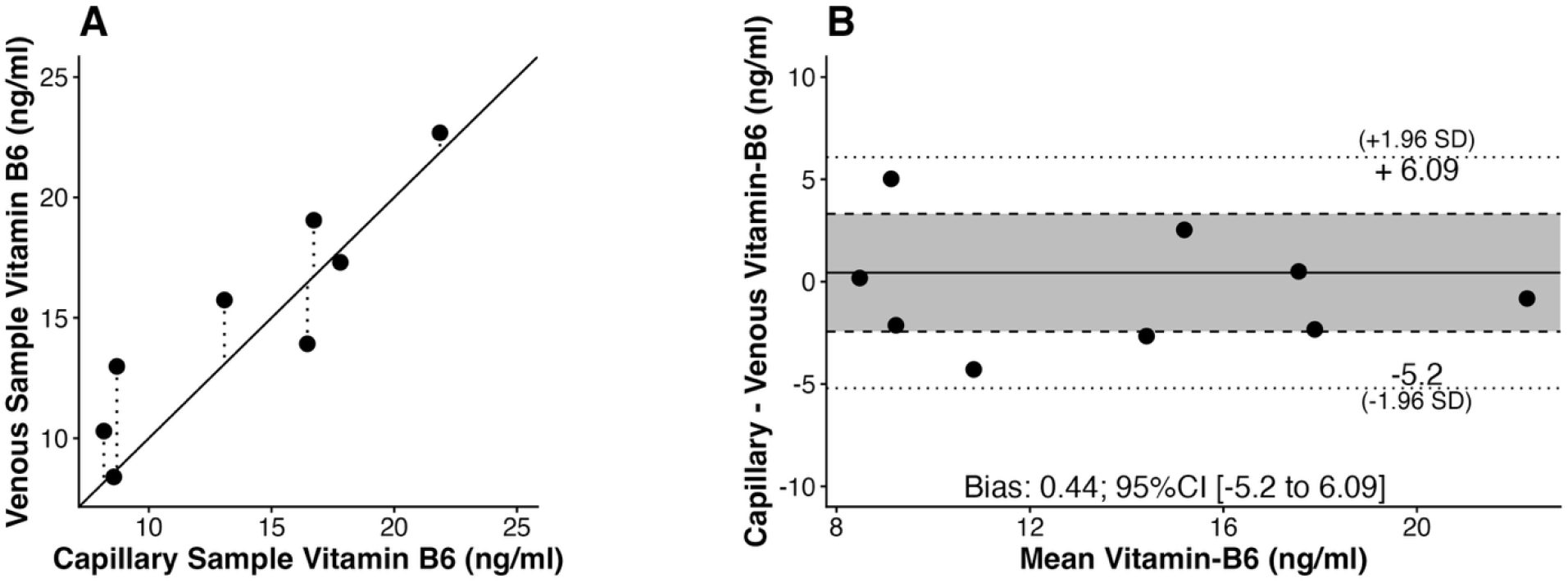
Agreement between venous and capillary Vitamin B6 measurements. (A) scatter plot showing the relationship between Vitamin B6 measurements from capillary samples collected using the TAP device and venous samples. Solid line represents the line of agreement (x=y). (B) Bland-Altman analysis plot of the difference between capillary and venous measurements against their mean. The solid line represents mean bias (0.44 ng/ml), and the dashed lines indicate the 95% limits of agreement (-5.20 to 6.09 ng/ml). Shaded regions illustrate the limits of agreement and confidence intervals.

**Figure 4.**
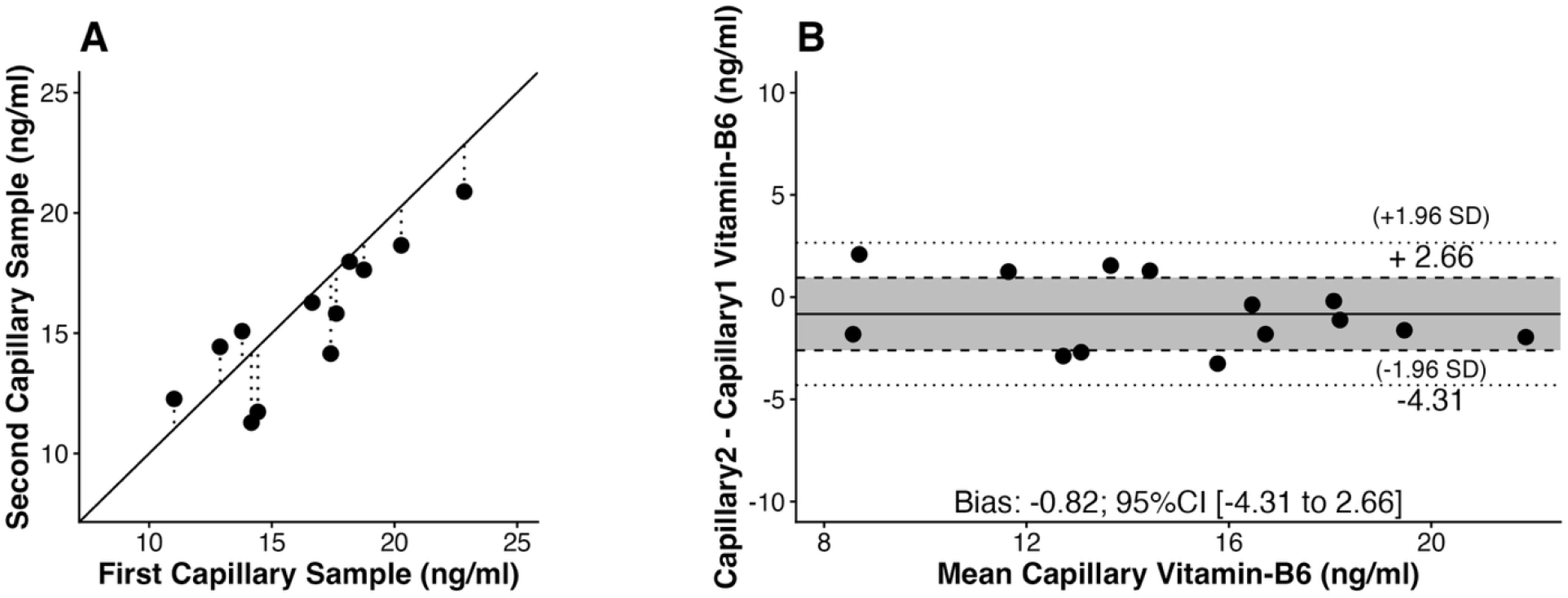
Reliability (repeatability) of capillary Vitamin B6 measurements. (A) scatter plot showing the relationship between repeated Vitamin B6 measurements from capillary samples. Solid line represents the line of agreement (x=y). (B) Bland-Altman analysis plot of the difference between repeated capillary measurements against their mean. The solid line represents mean bias (-0.82 ng/ml), and the dashed lines indicate the 95% limits of agreement (-4.31 to 2.66 ng/ml). Shaded regions illustrate the limits of agreement and confidence intervals.

### Is the concentration of Vitamin B6 in capillary blood comparable to that in venous blood?

Agreement between blood collected from the phlebotomy procedure (venous) and blood collected from the TAP procedure (capillary) for the assessment of Vitamin B6 concentration was assessed by Bland-Altman analysis. Bland-Altman analysis evaluates whether two methods can be used interchangeably by quantifying both systematic bias and random error (expressed as limits of agreement). Bland–Altman analysis revealed a mean bias of 0.44 ng/ml (SD 2.88), with 95% limits of agreement from −5.20 to 6.09 ng/ml. The standard deviation of the differences was 19.9% of the sample mean, which was comparable to previously reported within-person biological variation for Vitamin B6 (CVI = 20%, Talwar et al., 2005, Aarsand et al., 2026), indicating differences between methods are no greater than normal day□to□day physiological variation in Vitamin B6. There was no evidence of proportional bias (β = -0.078, 95% CI −0.61 to 0.45, p = 0.74), suggesting that the difference between capillary and venous Vitamin B6 measurements was consistent across the measurement range.

### Does the blood collected using the TAP capillary device provide a consistent measurement of Vitamin B6 concentration?

Repeated capillary samples produced highly consistent Vitamin B6 measurements. Reliability of Vitamin B6 measurements obtained using the TAP capillary device was assessed using Bland-Altman analysis, intraclass correlation, and measures of precision. Bland-Altman analysis showed no significant bias (Bias = -0.82 ng/ml, p = 0.106), with 95% limits of agreement from - 4.31 to 2.66 ng/ml. The standard deviation of the differences (1.78ng/ml) represented 12.4% of the sample mean, which is smaller in magnitude than previously reported within-person biological variation for Vitamin B6 (CVI = 20%, Talwar et al., 2005, Aarsand et al., 2026). The intraclass correlation coefficient indicated good to excellent reliability (ICC(A,1) = 0.888 [95%CI 0.680 to 0.963], p < 0.001). The absolute measurement error (SEM) was 1.39 ng/mL, and relative variability (CV) was 8.97%. Together, these results indicate that repeated capillary sampling from different parts of the capillary bed provides stable Vitamin B6 measurements, with low measurement error relative to biological variability.

### Was there a significant difference in plasma Vitamin B6 concentration between the autistic and non-autistic groups?

As the data met all the required assumptions for a parametric test, an independent samples t-test was conducted to investigate whether there was a significant difference in Vitamin B6 concentration between the autistic and non-autistic participants (Figure 5), which revealed that there was no statistically significant difference in Vitamin B6 concentration between the autistic and non-autistic participants (t(15) = -1.45, p = .169).

**Figure 5.**
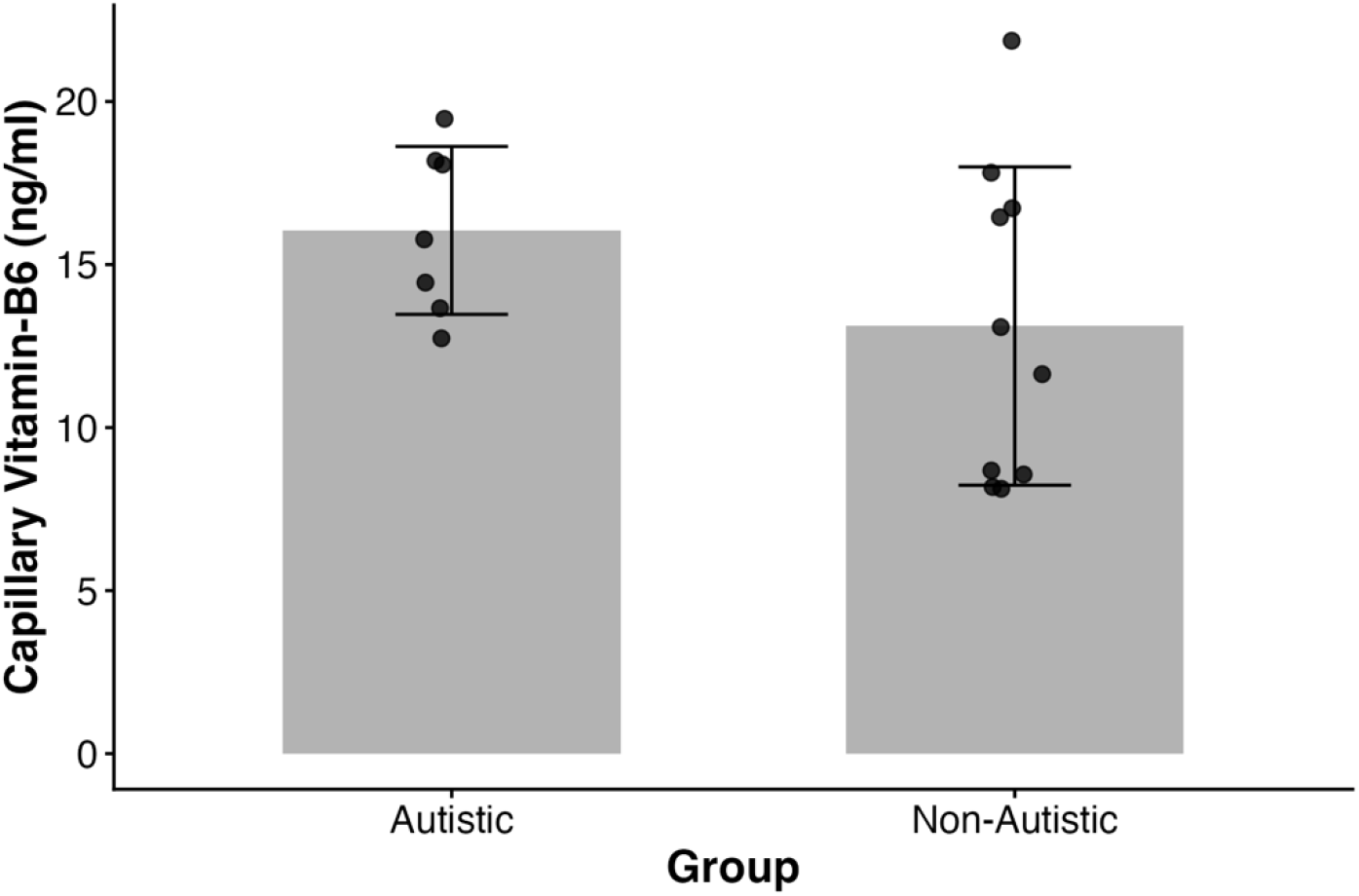
Comparing the total Vitamin B6 concentration between autistic and non-autistic individuals. Error bars reflect standard deviation.

## Discussion

We have replicated previous reports that drawing blood using the TAP device causes significantly less pain in non-autistic adults and extended this by showing that TAP device pain is comparably low in autistic adults. Furthermore, there was no significant difference in plasma total Vitamin B6 concentration between the autistic and non-autistic participants. Moreover, there was good agreement between capillary and venous blood assessment of concentration of Vitamin B6, indicating that future scientific studies of Vitamin B6 metabolism can analyse capillary blood and directly compare their results with studies that analyse venous blood. Finally, Vitamin B6 concentrations were consistent between the first and second capillary sample demonstrating strong reliability.

The data presented here strongly align with previous research conducted by Schröder et al. (2025), highlighting the tolerability of TAP devices. This will partly be due to the TAP device using an array of microneedles that only penetrate the upper layer of the skin, as opposed to phlebotomy, which must pass through the dermis into subcutaneous tissue. By avoiding the additional mechanical deformation of high-threshold mechanoreceptors that phlebotomy causes (Ialongo & Bernadini, 2016), TAP devices would induce less pain than phlebotomy. Furthermore, as Serafin et al. (2020) highlighted, needle size is directly correlated with pain in blood drawing procedures. As the TAP devices use an array of microneedles as opposed to a single large needle, this will result in less pain. This holds both potential research and clinical applications in providing a less painful technique for drawing blood. Finally, as Serra et al. (2018) highlighted, the large number of risks associated with phlebotomy may be mitigated by using TAP devices.

We predicted that the TAP devices would be significantly more painful for the autistic individuals than the non-autistic individuals, but in practice, we found no difference. However, there could be several features of our methodology that acted to reduce the difference. Firstly, participants were not screened for sensory hypo/hypersensitivity. Although the presence of sensory hypersensitivity is the most prevalent form of sensory difference in autistic adults, approximately 28.6% of autistic adults also report experiences of sensory hyposensitivity (MacLennan et al., 2022). It is possible that in our study, the individuals with hyposensitivity were overrepresented, as they would have been more likely to sign up for this type of research study. Furthermore, some of our efforts to recruit autistic participants may have reduced the anxiety that potentially underpins increased pain sensitivity. These efforts included sending videos of the procedure in advance of the session, along with a storyboard and pictures of the room. It is likely that by providing this information, participants experienced less intolerance of uncertainty and anxiety, which Jenkins et al. (2020) highlighted to be a prevalent challenge for autistic individuals. By reducing anxiety and intolerance of uncertainty effects, pain scores would likely be significantly lower, as Fischerauer et al. (2018) have previously identified.

We found no statistically significant differences in total Vitamin B6 concentration between the autistic and non-autistic groups, despite previous literature having reported that this is higher in autism (e.g. Adams et al., 2006). Nonetheless, the direction of difference we found was consistent with other studies that used the same Vitamin B6 measurement technique, and the non-significant finding is likely due to the small sample size that we recruited. Retrospective power analysis of Adams et al. (2006) indicates that a minimum of 16 participants would be required in each group to observe a significant difference with the effect sizes they reported. This is a critical limitation of this study as it was underpowered to detect the expected effect size that could be estimated from previous literature.

The present study demonstrates that capillary blood sampling using the TAP device provides valid and reliable measurement of Vitamin B6 concentration. Agreement analysis showed minimal bias between capillary and venous samples, with the variability of the differences comparable in magnitude to expected within-person biological variation. This indicates that disagreement between sampling methods is not greater than normal physiological fluctuation in Vitamin B6 levels. In addition, repeatability analyses showed high reliability across repeated capillary samples, with low measurement error and variability relative to biological variation. Together, these findings support the use of capillary blood sampling as a practical alternative to venous sampling for Vitamin B6 assessment, including repeated measures designs, particularly in studies requiring repeated or minimally invasive sampling.

This study had several limitations. Firstly, the sample size was smaller than suggested by a power analysis (see above), potentially resulting in fewer inferential statistics yielding significant results. But one of the main aims of this study was to investigate the feasibility of recruiting autistic individuals for a TAP blood drawing study; therefore, sample size was not considered to be a high priority. Moreover, phlebotomy was only conducted with the non-autistic participants, which did not allow for direct comparison of blood drawing techniques in the autistic group. Finally, future research investigating medical procedures, including blood draws, with autistic individuals should ensure measurements of sensory differences are taken to so that researchers can determine whether the nature of the study has produced a recruitment bias towards individuals who may be hypo- or hyper-sensitive.

In conclusion, this study highlights the benefits that alternative blood drawing techniques may have for autistic individuals, potentially improving research participation and reducing avoidance of medical appointments. By providing a technique that causes less discomfort than phlebotomy and which eliminates the small risk of damage to blood vessels associated with phlebotomy, TAP devices allow researchers to explore the metabolic and physiological differences underpinning autism by improving recruitment and widening participant diversity.

Concentrations of individual analytes should continue to be assessed between capillary and venous blood to create a more comprehensive understanding of the extent to which capillary blood samples can replace venous blood samples. Here, we have shown that for Vitamin B6, capillary samples are a valid and reliable alternative to venous phlebotomy. This study has provided a step towards more autism-friendly research practices and medical practices, enhancing the lives of participants, patients, researchers and clinicians.

## Data Availability

All data produced in the present study are available upon reasonable request to the authors

